# Predicting daily COVID-19 case rates from SARS-CoV-2 RNA concentrations across a diversity of wastewater catchments

**DOI:** 10.1101/2021.04.27.21256140

**Authors:** Alessandro Zulli, Annabelle Pan, Stephen M. Bart, Forrest W. Crawford, Edward H. Kaplan, Matthew Cartter, Albert I. Ko, Duncan Cozens, Marcela Sanchez, Doug E. Brackney, Jordan Peccia

## Abstract

We assessed the relationship between municipality COVID-19 case rates and SARS-CoV-2 concentrations in the primary sludge of corresponding wastewater treatment facilities. Over 1,000 daily primary sludge samples were collected from six wastewater treatment facilities with catchments serving 18 cities and towns in the State of Connecticut, USA. Samples were analyzed for SARS-CoV-2 RNA concentrations during a six-month time period that overlapped with fall 2020 and winter 2021 COVID-19 outbreaks in each municipality. We fit a single regression model to estimate reported case rates in the six municipalities from SARS-CoV-2 RNA concentrations collected daily from corresponding wastewater treatment facilities. Results demonstrate the ability of SARS-CoV-2 RNA concentrations in primary sludge to estimate COVID-19 reported case rates across treatment facilities and wastewater catchments, with coverage probabilities ranging from 0.94 to 0.96. Leave-one-out cross validation suggests that the model can be broadly applied to wastewater catchments that range in more than one order of magnitude in population served. Estimation of case rates from wastewater data can be useful in locations with limited testing availability or testing disparities, or delays in individual COVID-19 testing programs.

## Introduction

Wastewater surveillance has the potential to identify and track outbreaks of human pathogens that demonstrate gut tropism, including severe acute respiratory syndrome coronavirus 2 (SARS-CoV-2), the causative agent of COVID-19.^1^ Recent studies demonstrate that SARS-CoV-2 RNA concentrations in domestic wastewater reflect the rise and fall of COVID-19 cases based on daily positive tests in a community,^2-4^ and important epidemiological parameters such as the effective reproduction number can be estimated using wastewater SARS-CoV-2 concentrations, in addition to other epidemic indicators.^5^ If SARS-CoV-2 wastewater results are rapidly reported, they could provide a leading indicator of community infection rates over COVID-19 case rates and hospital admissions data.^5-7^

Estimates of infection based on COVID-19 case rates from testing of human specimens have been the standard for applying community interventions aimed at decreasing morbidity and mortality. Alternative approaches to COVID-19 testing are necessary to estimate the number of infections in locations with disparities in testing practices^8^, limited testing resources,^9^ or during outbreaks when testing capacity cannot meet demand. SARS-CoV-2 RNA concentration measurements in wastewater may resolve these issues, but quantitative relationships between SARS-CoV-2 concentration in wastewater and infection (or proxies of infection such as cases) are not well-resolved.

The following manuscript reports 192 days of monitoring daily primary sludge for SARS-CoV-2 concentrations across six different U.S. wastewater treatment facilities ranging in over one order of magnitude in flow rate, serving 18 municipalities and approximately 1 million residents in the State of Connecticut, USA. Regression models were evaluated to estimate community COVID-19 reported case rates from a time-course of primary sludge SARS-CoV-2 RNA concentrations.

## Materials and Methods

### Wastewater sampling

Primary sewage sludge samples of 40 to 45 ml in volume were collected daily from six wastewater treatment plants (WWTP) in the State of Connecticut, USA. **Table 1** lists the cities and towns served, and details the specific plant processes from which primary sludge was produced and samples were withdrawn. Grab samples from Stamford, Bridgeport, New Haven and New London were collected daily from August 3, 2020 to February 10, 2021. Samples from Hartford were collected daily from August 10, 2020 to February 10, 2021. Samples from Norwich were collected daily from August 17, 2020 to February 10, 2021. All samples were collected between 8 am and 9 am, stored at -20° C before being transported to Yale University laboratories on ice, and analyzed immediately upon arrival.

**Table 1.** Wastewater treatment plant characteristics

| WWTP Name | Municipalities served (total population) | Average flow rate (m<sup>3</sup>d<sup>-1</sup>) | Population served | Primary sludge process details |
| --- | --- | --- | --- | --- |
| Stamford | Stamford and Darien, CT (151,528) | 91,000 | 140,000 | Bar screens→primary clarifier→ de-gritting of primary sludge with hydrocyclones. Sample of thickened sludge collected after hydrocyclones. |
| Bridgeport West | Bridgeport, CT (144,900) | 120,000 | 105,000 | Bar screens→gravity thickener→primary clarifier. Sample collected at primary solids effluent pump. |
| New Haven | New Haven, Hamden, East Haven, Woodbridge, CT (228,862) | 151,000 | 200,000 | Bar screen→grit chamber→ primary clarifier→gravity thickener. Sample collected at effluent pump of gravity thickener. |
| Hartford, South Meadows | Hartford, West Hartford, Newington, Bloomfield, Wethersfield, CT (263,021) | 300,000 | 300,000 | Bar screen→grit chamber→ primary clarifier→gravity thickener. Sample collected at effluent pump of gravity thickener. |
| New London | New London and Waterford, CT (45,826) | 23,000 | 40,000 | Bar screen grit chamber→ primary sedimentation. Sample collected at primary solids pump. |
| Norwich | Norwich, Sprague, Franklin, Bozrah (46,495) | 32,000 | 22,500 | Bar screen→grit chamber→primary clarifier→gravity thickener. Sample collected at effluent pump from gravity thickener. |

### SARS-CoV-2 RNA primary sludge concentrations

To quantify SARS-CoV-2 RNA concentrations, the 40 to 45 ml sample was mixed on a vortexer for 1 minute and 0.5 ml of primary sludge was added to a commercial extraction kit optimized for the isolation of total RNA from raw wastewater (Zymo, *Quick*-RNA Fecal/Soil Microbe Microprep, wastewater protocol). Modifications to the extraction protocol included the addition of 0.1 ml of phenol/chloroform/isoamyl alcohol (25:24:1) in the initial bead beating step and eluting isolated RNA into 50 μl of ribonuclease-free water. Before September 20, 2020 and prior to the rapid rise in case rates in early October, primary sludge RNA extraction was accomplished using the RNeasy PowerSoil Total RNA kit (Qiagen) as previously described.^6^ For all RNA extracts, total RNA was measured by spectrophotometry and normalized to 200 ng μL^-1^ (NanoDrop, Thermo Fisher Scientific) prior to quantitative PCR to account for daily variations in sludge solids content.

SARS-CoV-2 RNA concentration was estimated using one-step qRT-PCR with SARS-CoV-2 N1 and N2 primer sets.^10, 11^ CrAssphage, a ubiquitous bacteriophage that is highly concentrated in the human gut,^12-14^ was quantified as a PCR control. All primer sets were run in separate reactions. Analysis was conducted using a one-step RT-qPCR kit (BioRad iTaq™ Universal Probes One-Step Kit). Triplicate 20 µL reactions using a 5x diluted template were run at 55° C for 10 min and 95° C for 1 min, followed by 40 cycles consisting of 95°C for 10 seconds and 55° C for 30 seconds^6^ and Ct values for triplicates were average. N1 and N2 primer set standards were constructed using ten-fold dilutions (5 × 10^1^ to 5 × 10^8^ copies per reaction) of the N gene transcripts for the SARS-CoV-2 Wuhan-Hu-1 strain. Concentrations in primary sludge samples were calculated using these standard curves and are presented as SARS-CoV-2 RNA copies per ng of total extracted RNA. Positive RNA and negative (no template) controls were included in all qRT-PCR runs. Less than 1.5% of samples were not available from the treatment facilities. In these cases, SARS-CoV-2 concentrations used in regression models were estimated by interpolating between the prior and subsequent measured SARS-CoV-2 concentration.

### COVID-19 test data for the cities served by the treatment facilities

The number of confirmed and probable COVID-19 cases per the Council of State and Territorial Epidemiologists case definition^15^ was provided by the Connecticut Department of Public Health (CT DPH) and used in this analysis. The date for each case was assigned, in order of preference, as test specimen collection date (if available), symptom onset date (if available), or date of report to the CT DPH. Greater than 90% of reported case values were by date of clinical specimen collection. Average test positivity rates over the study period were the following: Stamford (6.2%), Bridgeport (6.1%), New Haven (3.7%), Hartford (6.0%), New London (4.6%), Norwich (6.5%).

Cases were compiled from CT DPH data from the individual towns served by each wastewater treatment plant (**Table 1**) and adjusted per 100,000 population based on town census size. Under the assumption that each member of a population contributes an equivalent volumetric flow of raw wastewater and primary sludge across the cities and towns considered, the SARS-CoV-2 primary sludge concentrations reported here represent a per capita basis.

### Model development

Multiple regression analysis was used to fit COVID-19 reported cases per 100,000 population on the day of specimen collection to observed values of SARS-CoV-2 RNA copies per ng of total RNA in primary sludge. The model structure is

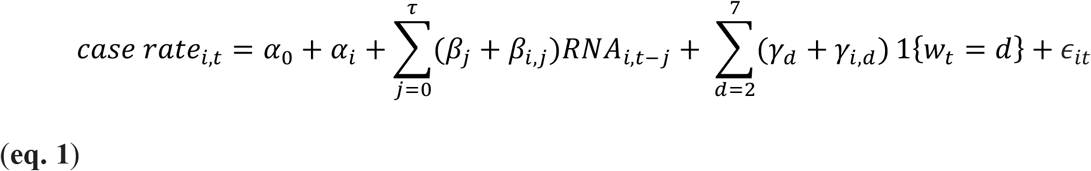

where the reported case rate is the number of new cases reported by date of specimen collection per 100,000 population, *i* represents municipality, *t* represents time (days), *α*_0_ is the model offset, *α*_*i*_ is the individual municipality model offset, *j* is the number of lagged days, *τ* is the maximum number of lagged days, *RNA*_*i,t*−*j*_ are RNA concentrations (SARS CoV-2 RNA copies ng^-1^ RNA), *w*_*t*_ indicates the day of the week on day *t, β*_*j*_, *β*_*i,j*_ and *γ*_*d*_ and *γ*_*i,d*_ are regression coefficients, *d* is day of the week (Friday is the reference day), and *ϵ*_*it*_ is the residual error associated with each observation. Day of the week addresses the known daily variations in testing behavior.^16^ Adjustment for population, which shows a strong collinearity with average treatment plant flow (simple linear regression: slope = 0.9 m^3^d^-1^ person^-1^, R^2^=0.93, p=0.002), was accomplished by considering the case rate per 100,000 residents as the dependent variable. A weighted least squares approach was used to weight (by reciprocal of the variance) case rate contributions to the model and reduce heteroskedasticity. A single model covering all treatment facilities was fit using the entire RNA concentration and case rate data set from all six treatment facilities from August 3, 2020 to February 10, 2021.

The statistical significance for predictive parameters was analyzed through a two-tailed t-test performed on the regression coefficients for all RNA concentration lags, and treatment plant and day of the week offsets. Akaike Information Criteria (AIC), Bayesian Information Criteria (BIC) F test, and coverage probability (calculated as the proportion of times that measured case rates fell within the 95% prediction interval of the model-estimated case rates), were used to select the maximum number of lagged RNA concentrations included in the model. A leave-one-out cross validation was utilized to test the model’s ability to estimate COVID-19 reported case rates for other wastewater catchments. Six regression models, similar to **Equation 1**, with the exception that municipality was not included, were trained by leaving out data from one of the six treatment facilities. These models were then utilized to estimate case rates in the cities covered by the treatment facility excluded from the model training set.

## Results

### SARS-CoV-2 concentrations

The SARS-CoV-2 RNA concentrations in primary sludge for the six treatment facilities are shown in **Figure 1** from August 3, 2020 to February 10, 2021 and reflect fall COVID-19 outbreaks in each city. The bottom row of **Figure 1** displays the reported case rates over the same time period in the municipalities served by the six treatment plants. The timing of SARS-CoV-2 RNA concentrations in primary sludge visually tracks the dynamics in reported case rates, with observed increases in primary sludge RNA concentrations at each plant coinciding with the start of the fall 2020 outbreaks for the corresponding cities.

**Figure 1.**
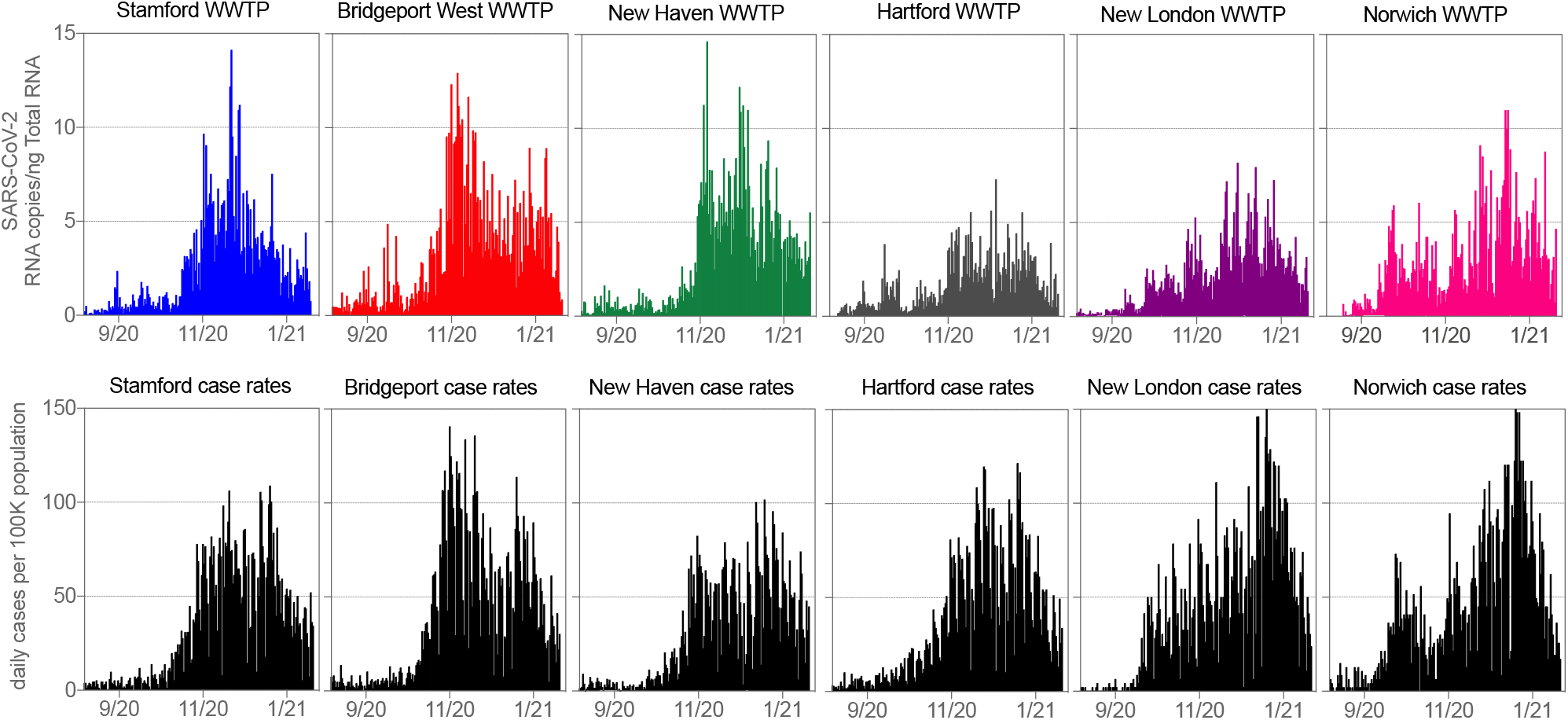
(top) Daily SARS-CoV-2 RNA concentrations in primary sludge of the six WWTP’s considered in this study. (bottom) Daily COVID-19 cases by date of specimen collection per 100,000 residents for the cities and towns served by the above wastewater treatment plants (**see Table 1**).

### Model selection and validation

This study estimated COVID-19 case rates using the regression model presented in equation 1 which included lagged SARS-CoV-2 RNA wastewater concentrations, municipality, and day of the week as variables. An initial analysis was conducted to determine the maximum number of prior days of wastewater RNA concentrations (*RNA*_*i,t*−*j*_, lagged days) to use. **Table 2** presents Akaike Information Criterion (AIC), Bayesian Information Criterion (BIC), F statistics with p values, and coverage probabilities for maximum lags ranging from 0 to 6 days and indicates optimal model fit with minimized model complexity when using lagged RNA concentrations from 0 to 4 days. Graphical comparisons of measured reported case rates versus model-estimated case rates are provided in **Figure 2** for each WWTP using models with lagged RNA concentrations from 0 to 4 days. When applied to the six individual municipalities, resulting coverage probabilities range was 0.93 to 0.96, with a root mean standard error of 13.1 cases per 100,000 population for the entire model (see **Figure S1** for model confidence intervals). **Tables S1-7** provide regression coefficients, intercepts, and their significance for the general linear models in **Equation 1**. Regression coefficients were typically greatest at lags 0 and 1 day.

**Table 2.** Resulting parameters for model selection to determine the maximum lagged RNA concentration.

| Maximum lag $\tau$ | AIC | BIC | F statistic (p value) | Coverage Probability |
| --- | --- | --- | --- | --- |
| 0 | 9835 | 10180 |  | 0.94 |
| 1 | 9663 | 10048 | 35.6 (<0.01) | 0.95 |
| 2 | 9613 | 10020 | 12.8 (<0.01) | 0.95 |
| 3 | 9557 | 10013 | 8.5 (<0.01) | 0.95 |
| 4 | 9543 | 9999 | 9.2 (<0.01) | 0.95 |
| 5 | 9521 | 10018 | 3.6 (<0.01) | 0.95 |
| 6 | 9536 | 10034 | 4.1 (<0.01) | 0.95 |

**Figure 2.**
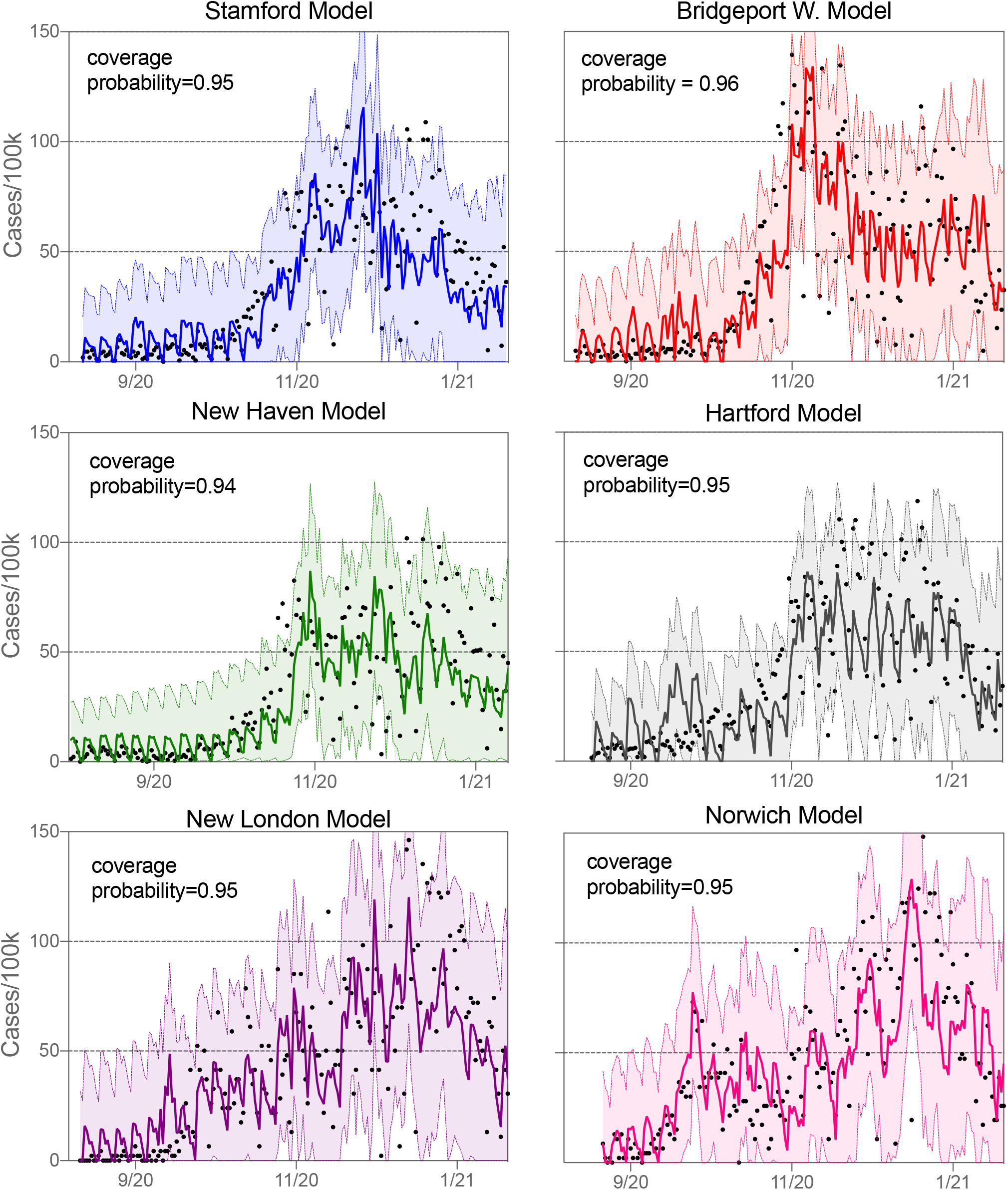
Comparison between COVID-19 case rates by day of clinical specimen collection and estimated case rates using RNA concentrations lagged from 0 to 4 days. The model is depicted by a solid line, measured cases are black dots, and 95% prediction intervals are shown by shading. *F*=37.42, *p*<0.001.

Results of the leave-one-out cross validation analysis revealed coverage probabilities (95% prediction intervals) of the estimated case rates that ranged from 0.90 to 0.98 for the six different municipalities considered, suggesting that the model can provide estimates of measured cases from sludge SARS-CoV-2 RNA concentrations for a variety of wastewater catchments that contain treatment facilities that produce primary sludge (**Table 3**).

**Table 3.**
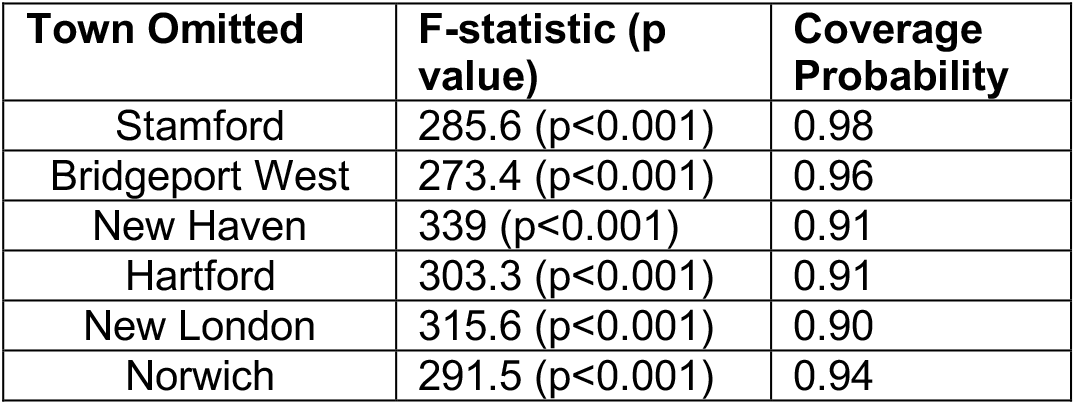
Leave-one-out cross validation results for the general model^1^

## Discussion

Estimating the number of infections directly from wastewater pathogen concentrations is a central goal of wastewater surveillance practice. The gut tropism of coronaviruses and widespread COVID-19 testing provide a unique opportunity to develop these tools. We monitored daily SARS-CoV-2 RNA concentrations in the primary sludge of six wastewater treatment plants that covered 18 U.S. municipalities and explored regression analyses to estimate COVID-19 cases rates on the day of specimen collection. Our results demonstrate the feasibility, utility, and simplicity of estimating COVID-19 case rates across a variety of different wastewater treatment catchments.

Prior studies have noted a concordance in wastewater SARS-CoV-2 RNA concentrations with other indicators of infection. Early work in the 2020 COVID-19 outbreak observed similar behavior between reported daily positive COVID-19 testing results in a community and the SARS-CoV-2 RNA concentrations in that community’s wastewater, ^17^ and relationships between wastewater RNA concentrations and COVID-19 reported cases have been reported for SARS-CoV-2 and polio virus. ^2, 7, 18, 19^ Several features of this study are unique to these prior studies and mark advances in the science underpinning wastewater-based epidemiology. We estimate COVID-19 case rates solely from current and prior days of primary sludge RNA concentrations and day of the week across treatment plants that range in more than one order of magnitude in size as measured by average flow rate and population served. Inclusion of concentration data from prior days yielded a model that can accounts for the previously observed offsets ^6, 7, 18^ between wastewater RNA concentrations and reported case data. The regression model used revealed that concentration lags of 0 to 1 day best predict case rates and confirms the previously described 0 to 2 day lag between case rates by date of specimen collection and SARS-CoV-2 concentrations in primary sludge^6^. A single general regression model trained by pooled plant data in a leave-one-out analysis was able to accurately estimate case rates for communities served by different domestic treatment plants, suggesting that this model could be extended more broadly to a variety of communities. In this case, treatment plants produced sludge using different primary treatment schemes and served populations that varied by more than an order of magnitude in size (**Table 1**). The approach for utilizing primary sludge instead of raw wastewater allows for rapid sampling of a mixed influent stream without the use of specialized sampling equipment. The concentration of SARS-CoV-2 in wastewater solids is greater than that in raw wastewater^3^ and negates the need for concentration steps often required when using untreated wastewater. The 0.5 ml sample volume considered in this study should enable automation of RNA extraction. SARS-CoV-2 concentrations are likely not impacted by precipitation events (infiltration and inflow) that might dilute concentrations of SARS-CoV-2 in the aqueous phase of raw wastewater.

The ability to estimate case rates from etiological agent concentrations in wastewater can be of significant epidemiological value. Case rates are a commonly used as a proxy for changes in community infection, and have been used as the standard for implementing non-pharmaceutical interventions and policies to reduce COVID-19 transmission and associated hospitalizations and deaths. In jurisdictions where testing is limited or does not exist, these models can be used as an independent estimate of infection rates. Wastewater RNA concentrations can be reported the same day the sample is collected, thus the statistical models used herein can be utilized to estimate up-to-date reported case rates in a community when COVID-19 testing data lags.

### Limitations

This modeling approach relies on statistical relationships between wastewater primary sludge RNA concentrations and COVID-19 case rates, which are primarily based on diagnostic test results. The reported prediction intervals reflect the variability in these reported case rates. While commonly used as a proxy, reported cases are believed to underestimate infection due to asymptomatic COVID-19 infections.^20^ Clinical testing volumes in the municipalities considered were dynamic over the study period; responding to events such as school openings, holidays, and shifting of resources to locations with increasing case rates. The measured RNA concentrations in wastewater are not subject to such variation in testing policy and should therefore exhibit a more direct relationship to the unobservable changes in SARS-CoV-2 infection in the community. While clinical testing data is considered an imperfect measure of infection, even in locations with strong testing programs, we note that the data resulting from testing programs has been indispensable in understanding the progression of outbreaks and initiating action to stem the spread of COVID-19 throughout the world. That case results can be estimated from SARS-CoV-2 concentrations in sewage sludge provides an added measure of confidence in the use of reported cases to monitor the epidemic.

### Summary

Measuring the concentration of pathogens in domestic wastewater can be a useful indicator of infection trends within a population. This study demonstrated that a single regression model populated by daily lagged SARS-CoV-2 sewage sludge concentrations could estimate COVID-19 case rates across communities served by six different wastewater treatment facilities. Cross-validation by leave-out analysis suggests the regression model can provide estimates of COVID-19 case rates for a broad variety of treatment facilities that produce primary sludge. Estimating case rates from wastewater pathogen concentrations can be useful in locations with limited or delayed COVID-19 testing programs or for infectious diseases where individual testing programs are not well-developed.

## Data Availability

COVID-19 case rate data was obtained from the CT department of health. Plots containing the case rate data and SARS-CoV-2 wastewater concentrations are available at: https://yalecovidwastewater.com/

## Acknowledgements

This project was supported by Cooperative Agreement no. 6NU50CK000524-01 from the Centers for Disease Control and Prevention using funds from the COVID-19 Paycheck Protection Program and Health Care Enhancement Act Response Activities. This activity was reviewed by CDC and was conducted consistent with applicable federal law and CDC policy. See e.g., 45 C.F.R. part 46.102(l)(2), 21 C.F.R. part 56; 42 U.S.C. 241(d); 5 U.S.C. 552a; 44 U.S.C. 3501 et seq. The findings and conclusions of this report are those of the author(s) and do not necessarily represent the official position of the Centers for Disease Control and Prevention.

We wish to thank the Stamford, Bridgeport West, New Haven, Hartford South Meadows, New London, and Norwich, CT wastewater treatment facilities for assistance in sampling and participation in the study.

## Supporting Information

General offset and coefficients for the regression model (Table S1), Stamford specific offset and coefficients for the regression model (Table S2), New Haven specific offset and coefficients for the regression model (Table S3), Hartford specific offset and coefficients for the regression model (Table S4), New London specific offset and coefficients for the regression model (Table S5), Norwich specific offset and coefficients for the regression model (Table S6).

## Notes

### Competing Interest Statement

The authors have declared no competing interest.

### Clinical Trial

This work did not result from a clinical trial. It is a comparison of wastewater concentrations with COVID-19 cases. The COVID-19 cases were obtained from publically available data. No human subjects were involved and all data is de-identified before being publically reported.

### Author Declarations

No IRB is required. The study used publically available COVID-19 cased data. All data is de-identified.

