## Supporting Information for "Predicting daily COVID-19 case rates from SARS-CoV-2 RNA concentrations across a diversity of wastewater catchments"

**Table S1.** General regression offset and coefficients, equation 1 model.

| **Coefficient** | **Value** | **Standard Error** | **Significance (<0.05 bold)** |
| --- | --- | --- | --- |
| α_0_ | -2.19 | 3.87 | 0.57 |
| β_0 (lag0)_ | 2.69 | 0.81 | **0.00085** |
| β_1 (lag1)_ | 3.51 | 0.83 | **0.00002** |
| β_2 (lag2)_ | 0.87 | 0.89 | 0.32 |
| β_3 (lag3)_ | 1.28 | 0.83 | 0.13 |
| β_4 (lag4)_ | 3.64 | 0.82 | **0.000009** |
| γ _(Monday)_ | 6.76 | 5.07 | 0.18 |
| γ _(Tuesday)_ | 9.34 | 5.04 | 0.06 |
| γ _(Wednesday)_ | 7.30 | 5.09 | 0.15 |
| γ _(Thursday)_ | 4.63 | 5.07 | 0.36 |
| γ _(Saturday)_ | -13.55 | 5.06 | **0.007** |
| γ _(Sunday)_ | -11.14 | 4.50 | **0.013** |

**Table S2**. Stamford specific regression offset and coefficients (Bridgeport is the reference group), equation 1 model.

| **Coefficient** | **Value** | **Standard Error** | **Significance (<0.05 bold)** |
| --- | --- | --- | --- |
| α_Stamford_ | 6.87 | 5.21 | 0.19 |
| β_0 (Stamford, lag0)_ | 0.87 | 1.26 | 0.50 |
| β_1 (Stamford, lag1)_ | -2.95 | 1.34 | **0.03** |
| β_2 (Stamford, lag2)_ | -0.53 | 1.41 | 0.70 |
| β_3 (Stamford, lag3)_ | 1.75 | 1.35 | 0.20 |
| β_4 (Stamford, lag4)_ | -0.35 | 1.27 | 0.78 |
| γ _(Stamford, Monday)_ | -2.68 | 6.87 | 0.70 |
| γ _(Stamford, Tuesday)_ | -6.01 | 6.84 | 0.38 |
| γ _(Stamford, Wednesday)_ | -5.57 | 6.91 | 0.42 |
| γ _(Stamford, Thursday)_ | -4.42 | 6.89 | 0.52 |
| γ _(Stamford, Saturday)_ | -0.53 | 6.86 | 0.93 |
| γ _(Stamford, Sunday)_ | 4.37 | 6.85 | 0.52 |

**Table S3.** New Haven specific offset and regression coefficients, equation 1 model.

| **Coefficient** | **Value** | **Standard Error** | **Significance (<0.05 bold)** |
| --- | --- | --- | --- |
| α_New Haven_ | 5.33 | 5.12 | 0.30 |
| β_0 (New Haven, lag0)_ | 0.44 | 1.24 | 0.72 |
| β_1 (New Haven, lag1)_ | -1.93 | 1.28 | 0.13 |
| β_2 (New Haven, lag2)_ | 0.50 | 1.31 | 0.70 |
| β_3 (New Haven, lag3)_ | -1.09 | 1.30 | 0.40 |
| β_4 (New Haven, lag4)_ | -2.28 | 1.26 | **0.07** |
| γ _(New Haven, Monday)_ | 0.67 | 6.81 | 0.92 |
| γ _(New Haven, Tuesday)_ | -2.78 | 6.75 | 0.68 |
| γ _(New Haven, Wednesday)_ | -1.28 | 6.75 | 0.85 |
| γ _(New Haven, Thursday)_ | -1.78 | 6.86 | 0.79 |
| γ _(New Haven, Saturday)_ | 2.69 | 6.80 | 0.69 |
| γ _(New Haven, Sunday)_ | 6.41 | 6.75 | 0.34 |

**Table S4.** Hartford specific regression offset and coefficients, equation 1 model.

| **Coefficient** | **Value** | **Standard Error** | **Significance (<0.05 bold)** |
| --- | --- | --- | --- |
| α_Hartford_ | -1.23 | 5.51 | 0.82 |
| β_0 (Hartford, lag0)_ | 3.06 | 1.67 | 0.06 |
| β_1 (Hartford, lag1)_ | 0.75 | 1.69 | 0.66 |
| β_2 (Hartford, lag2)_ | 3.92 | 1.73 | **0.02** |
| β_3 (Hartford, lag3)_ | 2.36 | 1.70 | 0.16 |
| β_4 (Hartford, lag4)_ | -0.52 | 1.69 | **0.75** |
| γ _(Hartford, Monday)_ | 3.47 | 7.05 | 0.62 |
| γ _(Hartford, Tuesday)_ | -0.93 | 7.00 | 0.89 |
| γ _(Hartford, Wednesday)_ | -1.08 | 7.07 | 0.87 |
| γ _(Hartford, Thursday)_ | -3.87 | 7.03 | 0.58 |
| γ _(Hartford, Saturday)_ | 1.56 | 6.95 | 0.82 |
| γ _(Hartford, Sunday)_ | 1.99 | 6.99 | 0.78 |

**Table S5.** New London specific regression offset and coefficients, equation 1 model.

| **Coefficient** | **Value** | **Standard Error** | **Significance (<0.05 bold)** |
| --- | --- | --- | --- |
| α_New London_ | 2.14 | 6.35 | 0.74 |
| β_0 (New London, lag0)_ | 3.99 | 1.88 | **0.03** |
| β_1 (New London, lag1)_ | -0.52 | 2.03 | 0.79 |
| β_2 (New London, lag2)_ | 0.42 | 2.10 | 0.84 |
| β_3 (New London, lag3)_ | 0.34 | 2.05 | 0.87 |
| β_4 (New London, lag4)_ | 1.08 | 1.91 | 0.57 |
| γ _(New London, Monday)_ | 4.89 | 8.13 | 0.55 |
| γ _(New London, Tuesday)_ | 1.39 | 8.17 | 0.86 |
| γ _(New London, Wednesday)_ | -2.07 | 8.15 | 0.80 |
| γ _(New London, Thursday)_ | 1.30 | 8.22 | 0.87 |
| γ _(New London, Saturday)_ | 0.99 | 8.09 | 0.90 |
| γ _(New London, Sunday)_ | 2.56 | 8.12 | 0.75 |

**Table S6.** Norwich specific regression offset and coefficients, equation 1 model.

| **Coefficient** | **Value** | **Standard Error** | **Significance (<0.05 bold)** |
| --- | --- | --- | --- |
| α_Norwich_ | -0.60 | 6.36 | 0.92 |
| β_0 (Norwich, lag0)_ | 0.75 | 1.41 | 0.60 |
| β_1 (Norwich, lag1)_ | -0.44 | 1.5 | 0.78 |
| β_2 (Norwich, lag2)_ | 2.10 | 1.58 | 0.18 |
| β_3 (Norwich, lag3)_ | -0.50 | 1.42 | 0.72 |
| β_4 (Norwich, lag4)_ | -0.018 | 1.32 | 0.98 |
| γ _(Norwich, Monday)_ | 2.95 | 8.11 | 0.72 |
| γ _(Norwich, Tuesday)_ | 1.28 | 8.04 | 0.87 |
| γ _(Norwich, Wednesday)_ | 4.42 | 8.03 | 0.58 |
| γ _(Norwich, Thursday)_ | 3.08 | 8.15 | 0.71 |
| γ _(Norwich, Saturday)_ | 5.86 | 8.01 | 0.46 |
| γ _(Norwich, Sunday)_ | 5.86 | 8.01 | 0.46 |

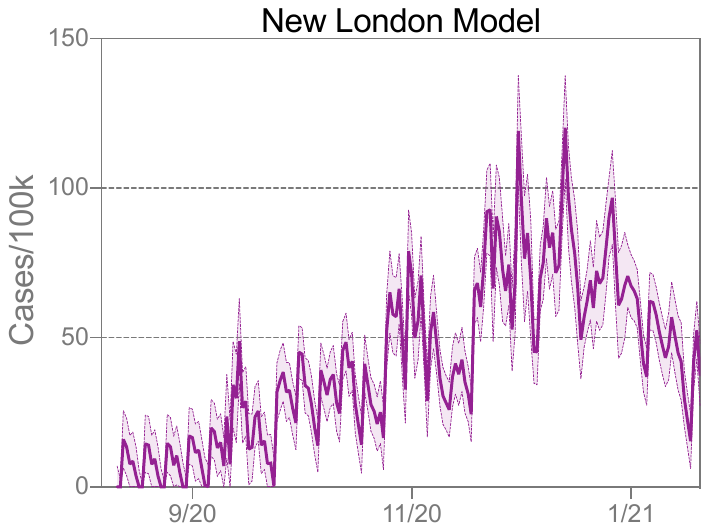

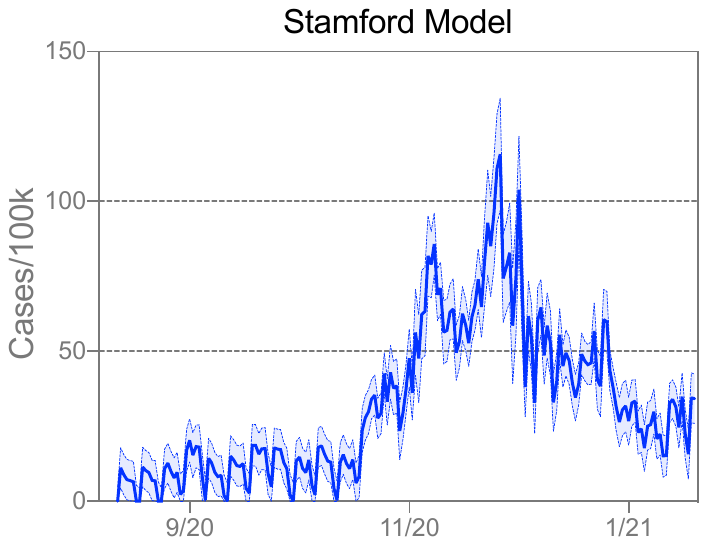

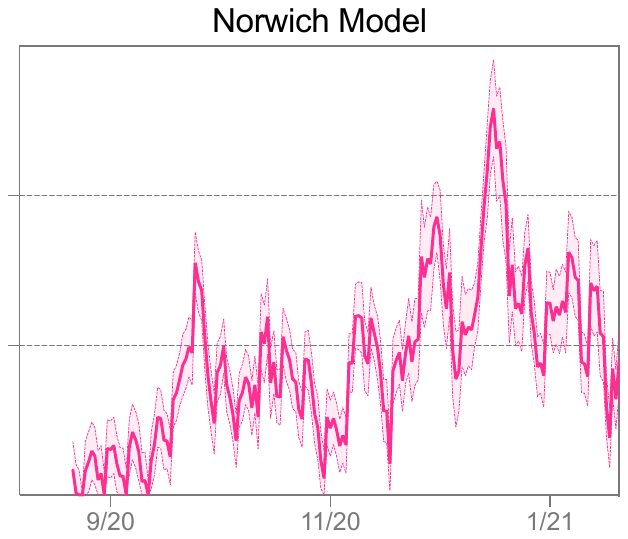

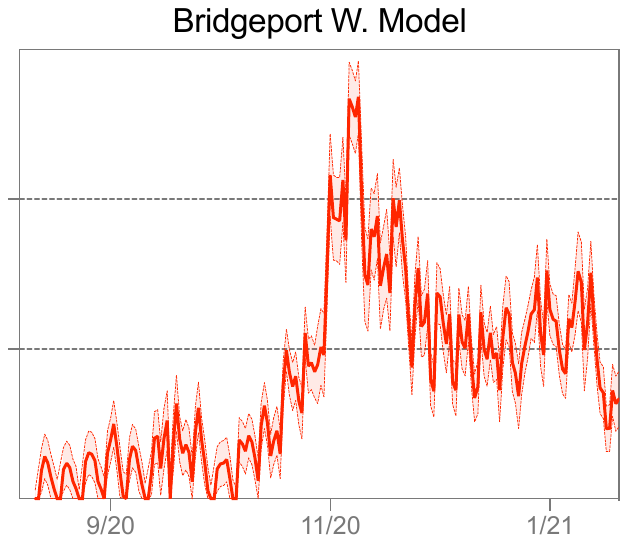

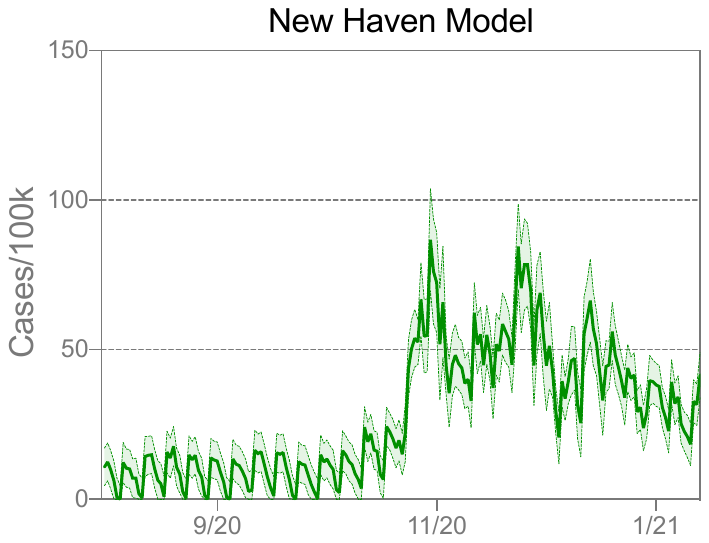

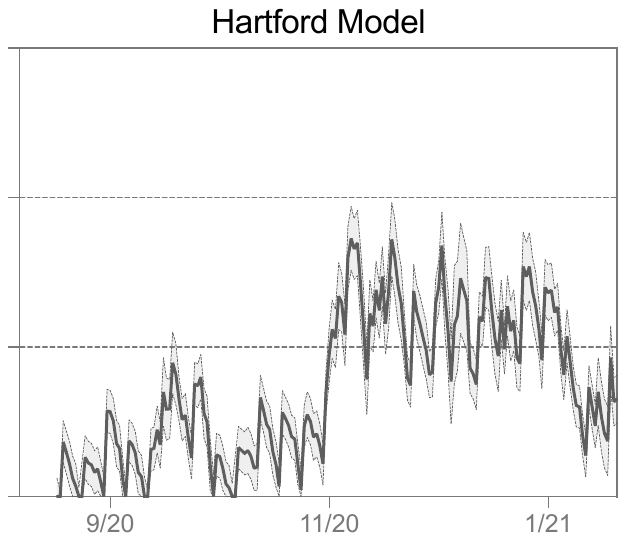

**Figure S1.** COVID-19 case rate model (solid line), depicting 95% confidence intervals (shaded areas). *F*=37.76, *p*<0.001.
